# Treatment Outcome of Bedaquiline-Based Oral Regimens for Drug-Resistant Tuberculosis in a High-Burden Setting

**DOI:** 10.1101/2025.09.01.25334877

**Authors:** Rashmi Ratnam, Parul Jain, Ram Awadh Singh Kushwaha, Amita Jain, Urmila Singh, Surya Kant

## Abstract

**Background:** In drug-resistant tuberculosis (DR-TB) management under India’s programmatic conditions, the World Health Organization (WHO) recommends two all-oral regimens for rifampicin-resistant (RR), multidrug-resistant (MDR), and extensively drug-resistant (XDR) TB: a longer M/XDR-TB regimen (18–20 months) and a shorter bedaquiline-containing MDR/RR-TB regimen (9–11 months). Their comparative effectiveness in India remains under-evaluated.

**Objective:** To compare clinical and microbiological outcomes of the longer versus shorter all-oral regimens in a prospective observational cohort.

**Methods:** In a prospective observational cohort study **f**rom January to December 2022, 906 newly diagnosed MDR-TB patients at a tertiary care center in Lucknow, India, were enrolled— 691 on the longer regimen and 215 on the shorter regimen. Six-month culture conversion and final treatment outcomes were recorded. Multivariable logistic regression adjusted for age, sex, baseline smear status, and comorbidities.

**Results:** At 6 months, culture conversion was higher with the longer regimen (79.8% vs 67.9%; absolute difference 11.9%; p<0.001; aOR 1.51, 95% CI 1.12–2.04). Favorable treatment outcomes also favored the longer regimen (77.9% vs 69.2%; 8.7%; p=0.010; aOR 1.37, 95% CI 1.05–1.79). In subgroup analyses, differences were significant in pulmonary TB and among women; pulmonary TB independently predicted favorable outcome (aOR 1.46, 95% CI 1.01– 2.13).

**Conclusions:** In this single-center cohort, the longer all-oral M/XDR-TB regimen yielded superior microbiological and clinical outcomes compared to the shorter regimen. These results support regimen selection tailored by patient profile and resistance pattern, and highlight the need for robust real-world tolerability and safety monitoring.

## Introduction

Tuberculosis (TB) remains a major global health challenge, particularly in low- and middle-income countries. Despite advances in diagnostics and treatment, drug-resistant TB (DR-TB) continues to impede control efforts, as evidenced by the 2022 WHO estimate of 450,000 new rifampicin-resistant/MDR-TB cases with only about 160,000 treated [1]. MDR-TB—resistant to at least rifampicin and isoniazid—is further complicated by the emergence of pre-extensively drug-resistant (Pre-XDR) and extensively drug-resistant (XDR) strains [2]. India bears a significant TB burden, accounting for approximately 27% of global cases and 26% of MDR/RR-TB cases, with a mortality rate of 30.8% in 2022 [3]. Moreover, resistance to newer agents like bedaquiline, delamanid, and linezolid is rising, jeopardizing recent therapeutic gains, especially as these drugs see expanded use under the national TB elimination program [4,5]. Patients Regimen:Longer Oral M/XDR-TB Regimen: Involves an 18-20 month treatment with Levofloxacin (Lfx), Bedaquiline (Bdq) for at least 6 months, Linezolid (Lzd), Clofazimine (Cfz), and Cycloserine (Cs).

Shorter Oral Bedaquiline:Containing MDR/RR-TB Regimen: A 9-11 month regimen, starting with Bdq, Lfx, Cfz, Pyrazinamide (Z), Ethambutol (E), high-dose Isoniazid (Hh), and Ethionamide (Eto) for 4-6 months, followed by Lfx, Cfz, Z, and E for 5 months.

To address DR-TB, the World Health Organization (WHO) recommends two standardized all-oral regimens: a longer regimen lasting 18–20 months and a shorter bedaquiline-containing regimen of 9–11 months duration [6]. The longer regimen includes three Group A drugs and one or more Group B drugs, offering around 80% efficacy but with challenges related to treatment duration and adherence [7]. The shorter regimen, structured into an intensive and continuation phase, is designed for improved adherence and cost-effectiveness, though it may carry a higher relapse risk in the presence of drug resistance [8]. While global studies have produced mixed findings on the non-inferiority of the shorter regimen [9], comparative data under India’s programmatic conditions remain limited. This study aims to address that gap by evaluating the clinical and microbiological outcomes of these regimens in a real-world Indian setting.

## Materials and Methods

### Study Population and Design

We conducted an observational prospective cohort study involving adult patients (≥18 years) diagnosed with rifampicin-resistant or multidrug-resistant tuberculosis (RR/MDR-TB), who were initiated on World Health Organization (WHO)-endorsed bedaquiline (BDQ)-based oral treatment regimens between January 2022 and December 2022 at our tertiary care center in North India having a drug resistance tuberculosis center (DR-TB) and an intermediate reference laboratory (IRL) for TB diagnostics under programmatic setting. Regimen selection was determined per programmatic guidelines and physician discretion, based on baseline drug susceptibility testing (DST) and patient-specific clinical conditions.

### Eligibility Criteria and Participant Selection

A total of 1,427 patients were screened, 906 met inclusion criteria (691 longer regimen; 215 shorter regimen).No formal sample size calculation was performed as this was an observational cohort study embedded within programmatic settings. All patients who met the inclusion criteria during the study period were consecutively enrolled to reduce selection bias.

Inclusion criteria comprised all patients with microbiologically confirmed RR/MDR-TB who received either the longer all-oral M/XDR-TB regimen or the shorter BDQ-containing MDR/RR-TB regimen under the national tuberculosis elimination program.

Exclusion criteria included (i) patients with incomplete baseline clinical or laboratory records, those transferred out before outcome documentation, and (iii) those not initiated on a standardized BDQ-based regimen (iv) those having severe extra-pulmonary TB (EP-TB) and milliary TB due to due to the complexity and severity of their clinical presentation.Eligible participants were consecutively enrolled from DRTB center to reduce selection bias.

### Minimization of Bias

Several strategies were employed to minimize bias. Selection bias was addressed by including all eligible patients consecutively over the defined study period. Misclassification bias was mitigated by cross-verifying treatment regimens from both patient treatment cards and pharmacy dispatch records. Measurement bias was reduced by standardizing data abstraction procedures and cross-checking culture conversion and outcome data against national TB reporting formats.

Additionally, potential confounding was minimized by stratifying results by key baseline variables (e.g., regimen type, resistance pattern, HIV status), and comparing outcomes across matched subgroups where applicable.

### Microbiological Investigation

Nucleic acid amplification testing (NAAT) was used as an upfront test for detecting rifampicin resistant tuberculosis. Samples were directly decontaminated using the NALC-NaOH method (0.5% NALC with 2% NaOH). Smear-positive samples were directly used for first-line and second-line drug resistance profiling using line probe assays (GenoTypeMTBDRplus and MTBDRsl v2.0, Hain Lifescience). Smear negative samples were cultured using the BD BACTEC MGIT 960 system. Liquid culture DST was also performed for isolates for moxifloxacin and linezolid. Cultured isolates were then used for indirect LPA. Resistance patterns were used to classify strains as MDR, pre-XDR, or XDR (by LC-DST) based on WHO guidelines.”

### Data Collection and Analysis

Baseline data included demographics, BMI, co-morbidities (diabetes, HIV), and TB site.Patients were followed up for clinical and microbiological outcomes until treatment completion. Clinical outcome was recorded at the end of treatment and was defined as favorable or unfavorable outcome. Favorable outcomes included cure or treatment completion, while unfavorable outcomes included treatment failure, death, loss to follow-up, or relapse. Treatment adherence was monitored using patient logs, and follow-up reports. Follow-up cultures were performed monthly as per NTEP guidelines. Patients with missing culture data beyond six months were excluded from the culture conversion analysis.

### Outcomes and Definitions

- *Microbiological Outcome:* Culture conversion at 6 months, (defined as two consecutive negative cultures at least 30 days apart).
- *Clinical Outcome:* Classified as; Favorable = cured/treatment completed; Unfavorable = treatment failure, death, loss to follow-up (LTFU) at the end of the treatment regimen.

### Statistical Analysis

Data were analyzed using SPSS v24. Continuous variables were assessed for normality using the Shapiro-Wilk test. Normally distributed data were compared using independent t-tests; otherwise, Mann-Whitney U tests were applied. Categorical variables were compared using chi-square tests. A Multivariable models were adjusted for potential confounders identified a priori, including age, sex, BMI, site of TB, HIV status, and diabetes. Odds ratios (OR) and 95% confidence intervals (CI) were reported.

We conducted a complete-case analysis: patients missing primary outcome data (interim culture conversion or final treatment outcome) were excluded from the corresponding analyses. No data imputation or sensitivity analyses were performed, given the low rate of missingness (≤3.3%) and the consistency of results across prespecified subgroups.

### Ethical Approval

The study was approved by the K. G. M. U. Institutional Ethics Committee (Ref No: 126th ECMIIB-Ph.D/P1).

## Results

### Patient Enrollment and Baseline Characteristics

A total of 1,427 patients were screened during the study period, of whom 906 met the inclusion criteria and were enrolled. Of these, 691 (76.3%) received the longer oral M/XDR-TB regimen and 215 (23.7%) received the shorter bedaquiline-containing MDR/RR-TB regimen. The enrollment flow is depicted in **Figure 1**.

**Figure 1.**
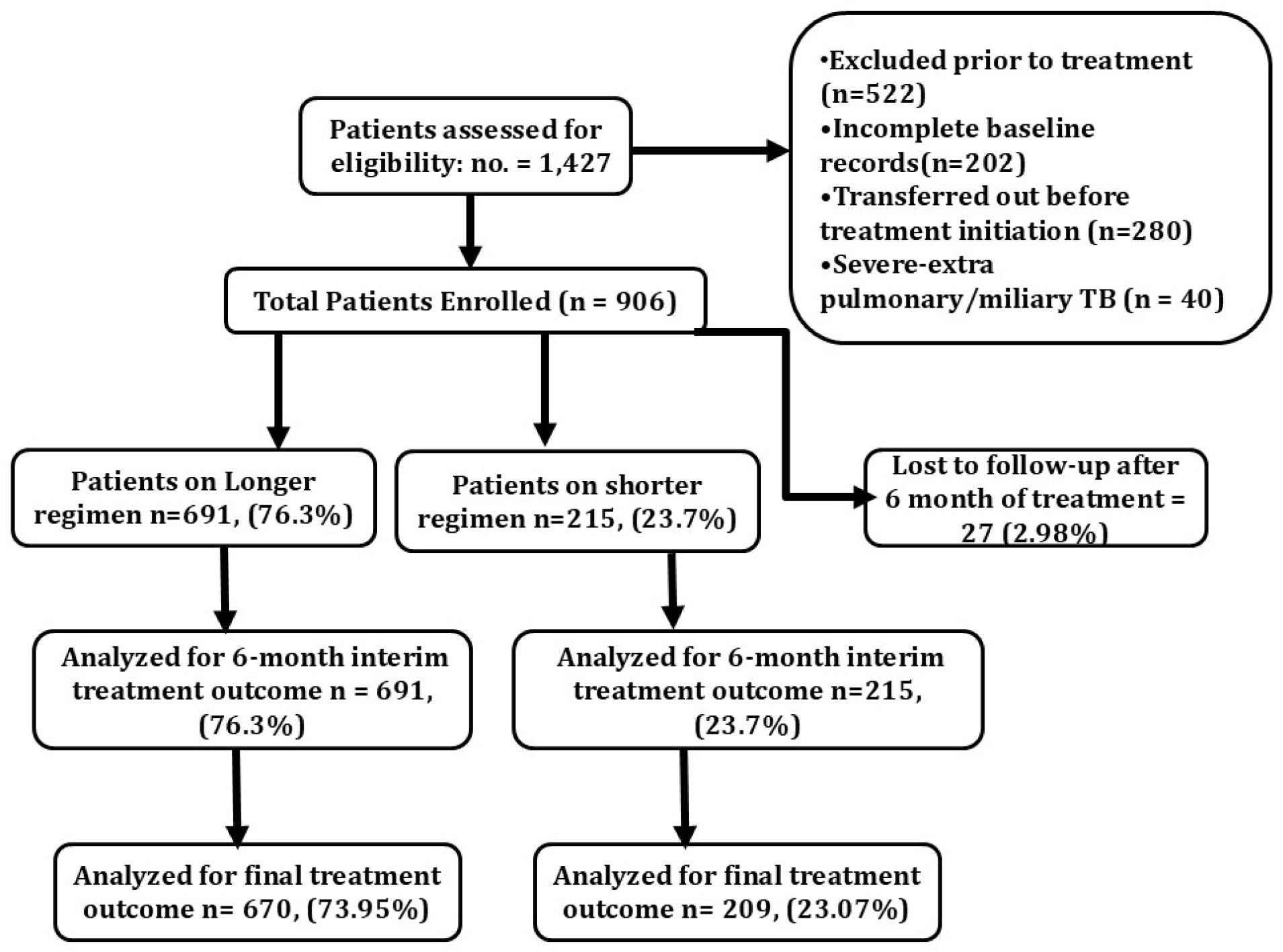
Flowchart showing patient screening, inclusion, exclusion, and treatment group allocation.

The mean age of enrolled patients was 42 ± 14 years, with males accounting for 63%. Approximately 28% of patients had a BMI <18.5 kg/m^2^, consistent with the high prevalence of malnutrition in TB. Baseline characteristics, including age, sex, BMI, HIV status, diabetes, and TB disease site, did not differ significantly between the two treatment groups as shown in **Table 1**.

**Table 1.** Baseline Demographic and Clinical Characteristics of Patients Receiving Longer and Shorter Regimens.

| Characteristics | Longer Oral Regimen (n=691) | Shorter Oral Regimen (n=215) | p Value |
| --- | --- | --- | --- |
| Gender |  |  |  |
| Male | 400 | 131 | 0.42 |
| Female | 291 | 83 |  |
| Age (in years) |  |  |  |
| <18 | 66 | 32 | 0.03 |
| >18 | 625 | 182 |  |
| BMI (mean ± SD) | 17.54 ± 7.54 | 17.46 ± 6.33 | 0.67 |
| Drug Resistance |  |  |  |
| MDR | 47 | 12 | 1 |
| Pre- XDR | 644 | 202 |  |
| Co-morbidities |  |  |  |
| Diabetic | 36 | 7 | 0.27 |
| Non- Diabetic | 655 | 208 |  |
| HIV Reactive | 4 | 2 | 0.63 |
| HIV Non Reactive | 687 | 212 |  |
| Site Of Diseases |  |  |  |
| Pulmonary | 630 | 193 | 0.59 |
| Extra Pulmonary | 61 | 22 |  |

Of the 906 patients enrolled, 19 (2.1%) had at least one missing baseline variable; the number and proportion of missing observations for each variable are detailed in Supplemental Table S1. Mean follow-up duration was 10.2 months (SD 2.1; range 3–18 months), with no significant difference between the longer and shorter regimen groups

### Microbiological and Clinical Outcomes

At 6 months, the culture conversion rate was significantly higher among patients on the longer regimen (79.8%, n = 579) compared to the shorter regimen (67.9%, n = 146; p = 0.0003). Favorable clinical outcomes were observed in 77.9% (n = 525) of the longer regimen group and 69.2% (n = 144) of the shorter regimen group (p = 0.010). These outcomes are summarized graphically in **Figure 2**.

**Figure 2.**
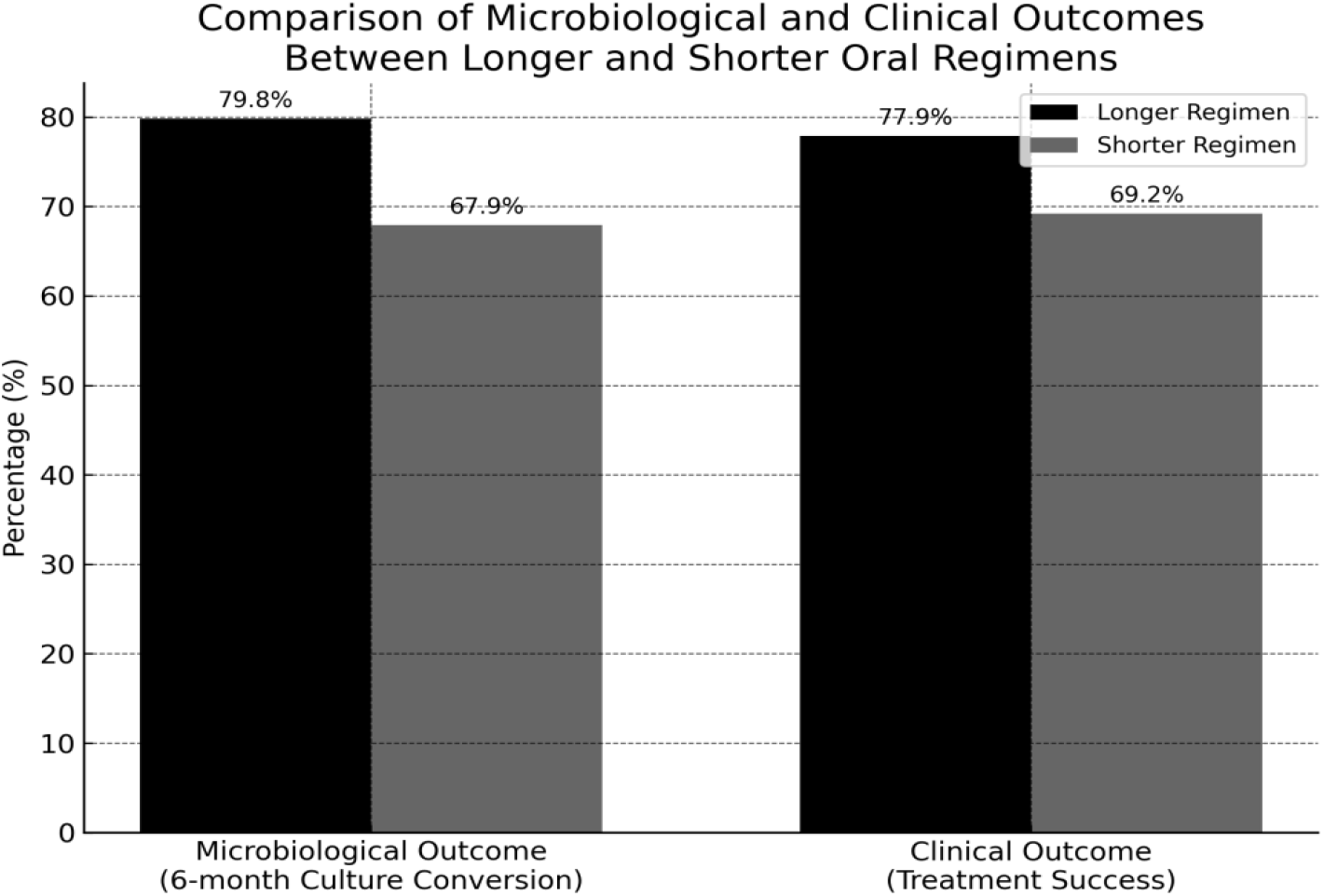
Comparative Bar Plot of Microbiological and Clinical Outcomes Between Longer and Shorter Oral Regimens.

### Subgroup Analysis

Patients with pulmonary TB showed significantly better outcomes on the longer regimen compared to the shorter regimen (p = 0.008). No significant differences were observed among patients with extrapulmonary TB (p = 1.000), diabetes (p = 0.560), HIV-positive status (p = 0.400), or low BMI. Interestingly, female patients on the longer regimen had significantly better outcomes than those on the shorter regimen (p = 0.040), whereas differences among male patients were not statistically significant (p = 0.180). These detailed subgroup comparisons are presented in **Tables 2a and 2b**.

**Table 2a.** Clinical Outcomes – Overall and Pulmonary vs Extra-pulmonary TB

| Subgroup | Regimen Type | Favorable | Unfavorable | p-value |
| --- | --- | --- | --- | --- |
| **All Patients** | Longer | 525 | 149 | 0.010 |
|  | Shorter | 144 | 64 |  |
| **Pulmonary TB** | Longer | 482 | 133 | 0.008 |
|  | Shorter | 130 | 59 |  |
| **Extra-pulmonary TB** | Longer | 44 | 16 | 1.000 |
|  | Shorter | 14 | 4 |  |

**Table 2b.** Clinical Outcomes – By Co-morbidities and Sex

| Subgroup | Regimen Type | Favorable | Unfavorable | p-value |
| --- | --- | --- | --- | --- |
| **Diabetic Patients** | Longer | 27 | 5 | 0.560 |
|  | Shorter | 7 | 0 |  |
| **HIV Positive Patients** | Longer | 1 | 0 | 0.400 |
|  | Shorter | 2 | 3 |  |
| **Female** | Longer | 224 | 60 | 0.040 |
|  | Shorter | 55 | 26 |  |
| **Male** | Longer | 301 | 89 | 0.18 |
|  | Shorter | 89 | 36 |  |

### Multivariable Logistic Regression Analysis

In adjusted analyses, patients receiving the longer regimen had significantly higher odds of a favorable clinical outcome (aOR = 1.51; 95% CI: 1.12–2.04; **p** = 0.006). Pulmonary TB was also independently associated with improved outcomes (aOR = 1.46; 95% CI: 1.01–2.13; **p** = 0.042). Other covariates—including age, sex, low BMI, diabetes, and HIV—were not significantly associated with treatment success. Full regression results are presented in **Table 3**, with key predictors visualized in **Figure 3**.

**Table 3.** Multivariable Logistic Regression for Predictors of Favorable Treatment Outcome

| Variable | Adjusted OR (aOR) | 95% CI | p-value |
| --- | --- | --- | --- |
| **Regimen (Longer)** | 1.51 | 1.12–2.04 | 0.006 |
| **Age >18 yrs** | 1.04 | 0.72–1.51 | 0.830 |
| **Male** | 1.09 | 0.83–1.43 | 0.520 |
| **BMI < 18.5** | 0.91 | 0.68–1.21 | 0.530 |
| **Pulmonary TB** | 1.46 | 1.01–2.13 | 0.042 |
| **Diabetic** | 0.96 | 0.56–1.63 | 0.880 |
| **HIV Positive** | 0.75 | 0.23–2.43 | 0.630 |

**Figure 3.**
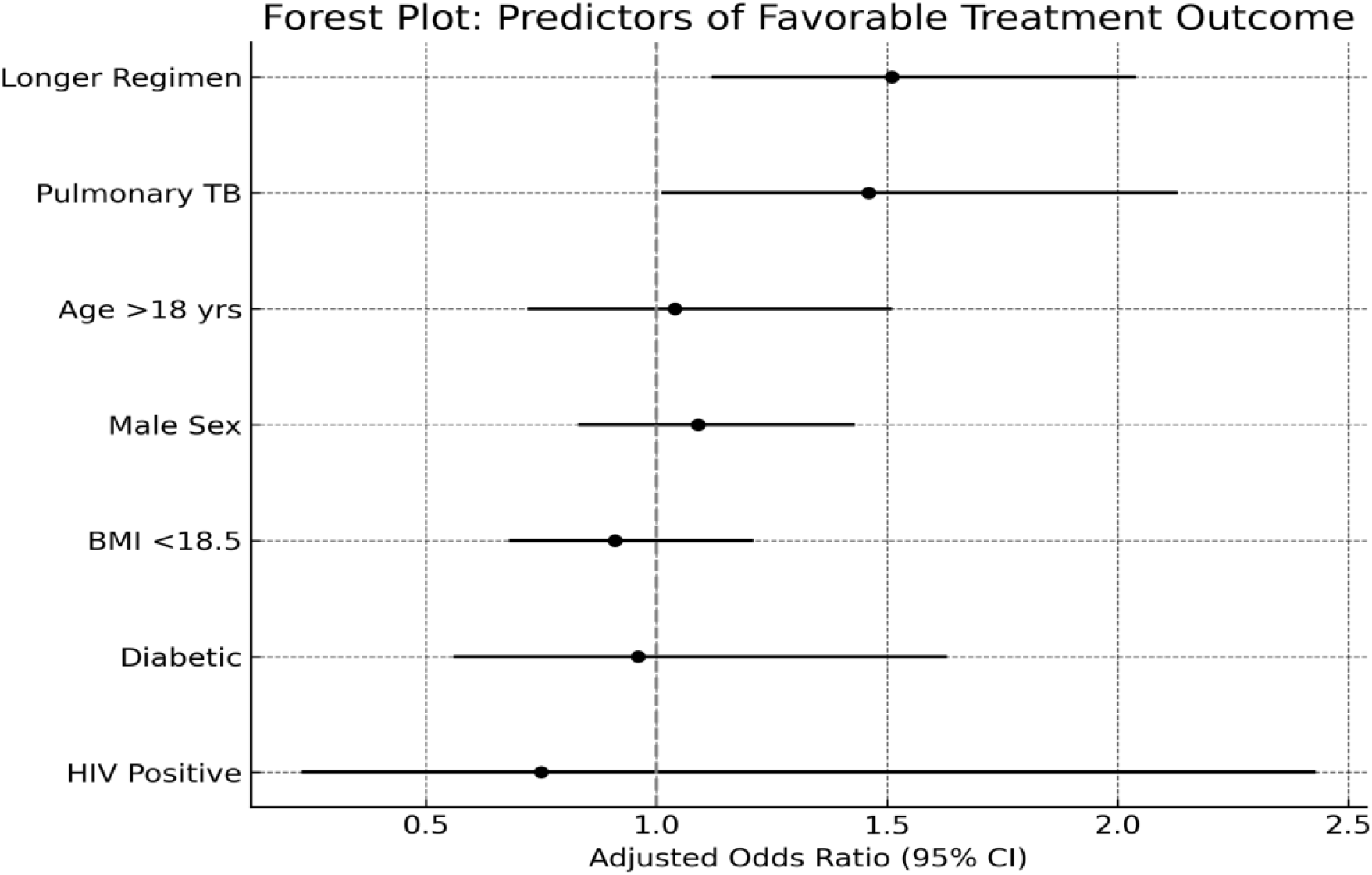
Forest plot displaying adjusted odds ratios and 95% confidence intervals from multivariable logistic regression, highlighting regimen type and pulmonary TB as independent predictors of favorable treatment outcomes.

## Discussion

The findings of this study underscore the importance of selecting appropriate regimens for managing MDR, pre-XDR, and XDR TB, particularly in high-burden settings like India. The longer all-oral regimen demonstrated superior clinical and microbiological outcomes compared to the shorter Bdq-containing regimen particularly in treating pulmonary TB, which is the most common and infectious form of the disease. These results are consistent with other studies that have emphasized the advantages of longer treatment regimens in achieving higher rates of culture conversion and preventing relapse due to their comprehensive drug coverage and reduced risk of resistance development, particularly in complex MDR/XDR TB cases [10, 11]. The superior outcomes observed with the longer regimen may be explained by its broader drug coverage and inclusion of drugs with potent sterilizing activity, such as linezolid and clofazimine, which have shown significant bactericidal and sterilizing activity against TB [12, 13], a critical phenomenon in preventing the transmission of drug-resistant strains. Studies from South Africa and other high-burden countries have similarly reported that longer regimens incorporating these drugs result in higher cure rates and lower mortality compared to shorter regimens [14, 15]. However, the prolonged treatment duration poses challenges related to patient adherence and potential adverse effects, necessitating careful patient monitoring and support [16, 17]. One limitation of our study is the lack of systematic data on treatment-emergent adverse events. Adverse drug reactions, particularly those associated with linezolid (e.g., myelosuppression, neuropathy) and clofazimine (e.g., hyperpigmentation, gastrointestinal intolerance), may affect adherence and outcomes. Future prospective studies should incorporate pharmacovigilance data to better characterize regimen tolerability.

The lower conversion rate indicates that while the shorter regimen is effective for many patients and being advantageous in terms of patient adherence and cost-effectiveness, it may not be as robust as the longer regimen in achieving early microbiological clearance. This aligns with previous findings that shorter regimens may be more susceptible to resistance-related treatment failures particularly in the presence of resistance to key drugs like fluoroquinolones and bedaquiline [18, 19], underscoring the importance of comprehensive drug susceptibility testing (DST) prior to regimen selection [20, 21, 22].

Given the high burden of MDR and XDR TB in India, optimizing treatment regimens based on local resistance patterns, patient demographics, and healthcare resources is crucial. The choice of regimen must balance efficacy, patient adherence, and healthcare system capacity. Additionally, more intensive patient management and monitoring due to the prolonged duration of treatment may result in better long-term outcomes and reduced transmission rates [23]. Further research should focus on refining shorter regimens to enhance their efficacy while maintaining adherence benefits, potentially through the inclusion of newer TB drugs and tailored treatment strategies [24, 25].

## Future Directions

To optimize TB management strategies, especially for drug-resistant forms, there is a need for continued research into newer drugs and treatment combinations. Recent studies have explored the potential of novel TB drugs like pretomanid and delamanid, either alone or in combination with bedaquiline, in shortening treatment duration while maintaining high efficacy rates [26, 27]. Additionally, the integration of molecular diagnostics and rapid DST into routine care could help in the early identification of resistance patterns and guide more effective treatment choices [28, 29].A study by the TB CARE consortium demonstrated that integrating operational research into national TB programs could enhance regimen effectiveness, improve patient outcomes, and support the adoption of evidence-based practices [30]. Though the present study did not show any impact of diabetes or HIV on treatment outcome, given the diverse demographic and clinical characteristics of TB patients in India, further studies should also explore the impact of comorbidities, such as diabetes and HIV, on treatment outcomes with different regimens [31,32].The success of TB control programs in countries like Peru and Russia, where individualized treatment regimens have been tailored based on comprehensive DST and patient-centered care models, provides valuable lessons for other high-burden settings [33, 34]. Policymakers should prioritize the availability of all essential TB drugs, strengthen diagnostic capacities, and ensure adequate funding for TB control programs.

This study adhered to the STROBE (Strengthening the Reporting of Observational Studies in Epidemiology) guidelines for reporting observational cohort studies. A completed STROBE checklist is provided as supplementary material.

## Limitations of the Study

While the study provides valuable insights, several limitations should be acknowledged. The observational design and single-center hospital based setting may limit the generalizability of the findings. Selection bias is also a potential issue, as patients were not randomized to treatment groups. The non-randomized design and clinical discretion in regimen allocation may have introduced selection bias and unmeasured confounding.. Propensity score matching or inverse probability weighting could be considered in future analyses to mitigate this bias. Moreover, the study did not account for all potential confounding factors, including baseline disease severity, duration of previous TB treatment, exact timing of culture conversion, and drug availability. Additionally, adherence was monitored via patient logs but not validated with objective adherence tools such as digital monitors or DOT.

## Conclusion

This study adds to growing evidence favoring longer, all-oral regimens. These regimens may be more effective for managing complex drug-resistant TB cases in high-burden settings. However, balancing the benefits of shorter regimens for patient adherence and cost with the need for higher efficacy remains a critical challenge. Future research should continue to explore innovative treatment strategies, optimize regimen composition using newer agents, and integrate individualized treatment monitoring to enhance outcomes in drug-resistant TB.

## Data Availability

All relevant data underlying the findings of this study are contained within the manuscript and its Supporting Information files. De-identified patient-level data used for analysis are available from the corresponding author upon reasonable request, subject to approval from the Institutional Ethics Committee of King George's Medical University, Lucknow, India, to ensure compliance with patient confidentiality requirements.

## Acknowledgments

We thank the staff of Intermediate reference laboratory King Georges Medical University and National Tuberculosis Elimination Progamme (NTEP) for kits and consumables.

## Legends

**Supplemental Table S1.** (baseline missingness):

| Variable | Missing, n (%) |
| --- | --- |
| Age (years) | 0 (0.0) |
| Sex | 0 (0.0) |
| Body mass index (BMI, kg/m<sup>2</sup>) | 0 (0.0) |
| Baseline sputum smear status | 0 (0.0) |
| HIV status | 0 (0.0) |
| Diabetes mellitus | 0 (0.0) |
| Chest X-ray cavitation | 0 (0.0) |
| Prior TB treatment history | 0 (0.0) |
| Any other comorbidity | 0 (0.0) |

**Note: All baseline variables were complete because patients with any missing baseline data (n=522) were excluded prior to analysis.**

## S1 Checklist. STROBE Checklist for Cohort Study

This checklist provides the page and section references in the manuscript **“Treatment Outcome of Bedaquiline-Based Oral Regimens for Drug-Resistant Tuberculosis in a High-Burden Setting”** that correspond to the STROBE recommendations for reporting cohort studies. This file is provided as Supporting Information according to PLOS Global Public Health submission guidelines.

## STROBE Statement—Checklist for Cohort Study

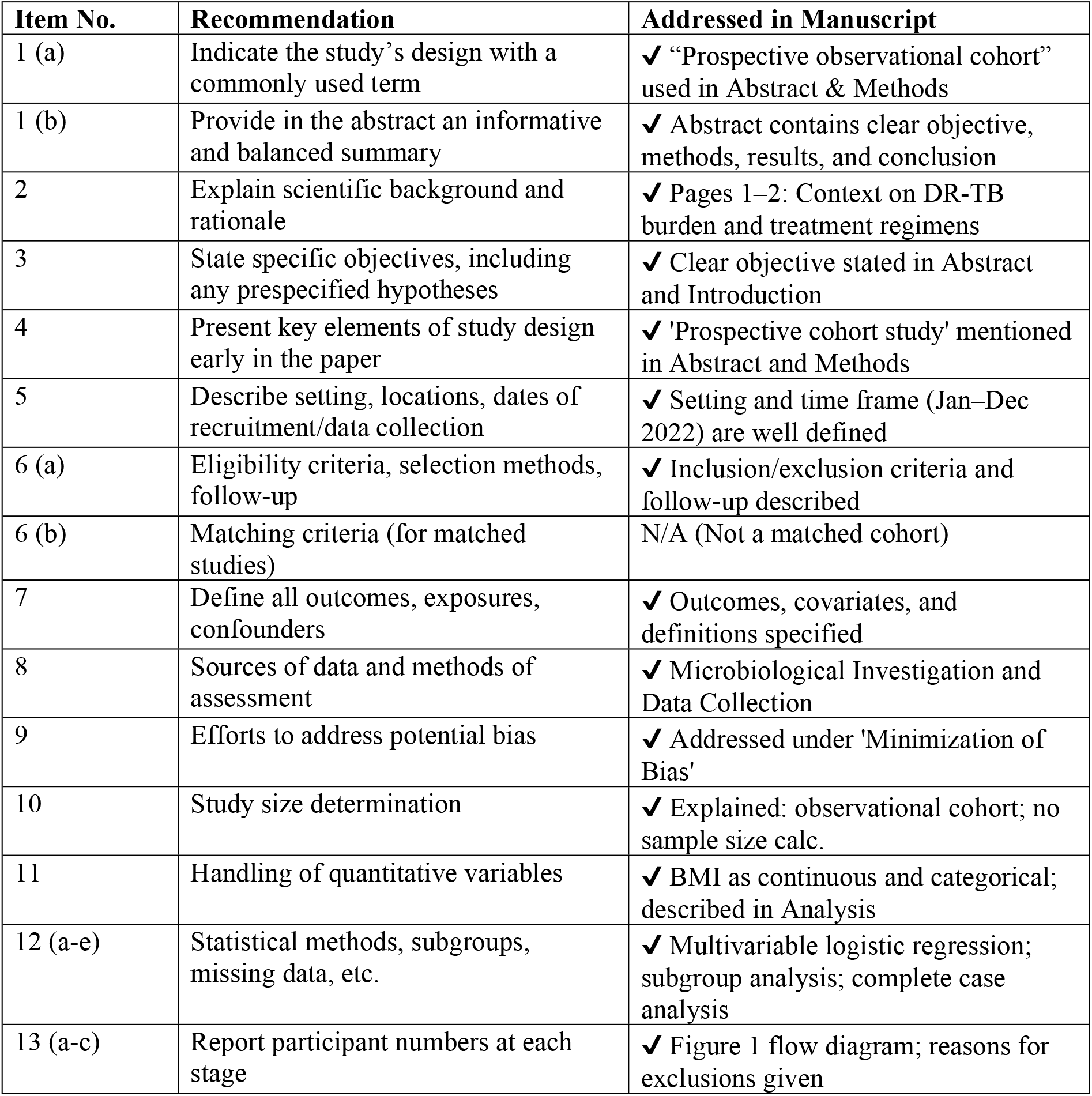
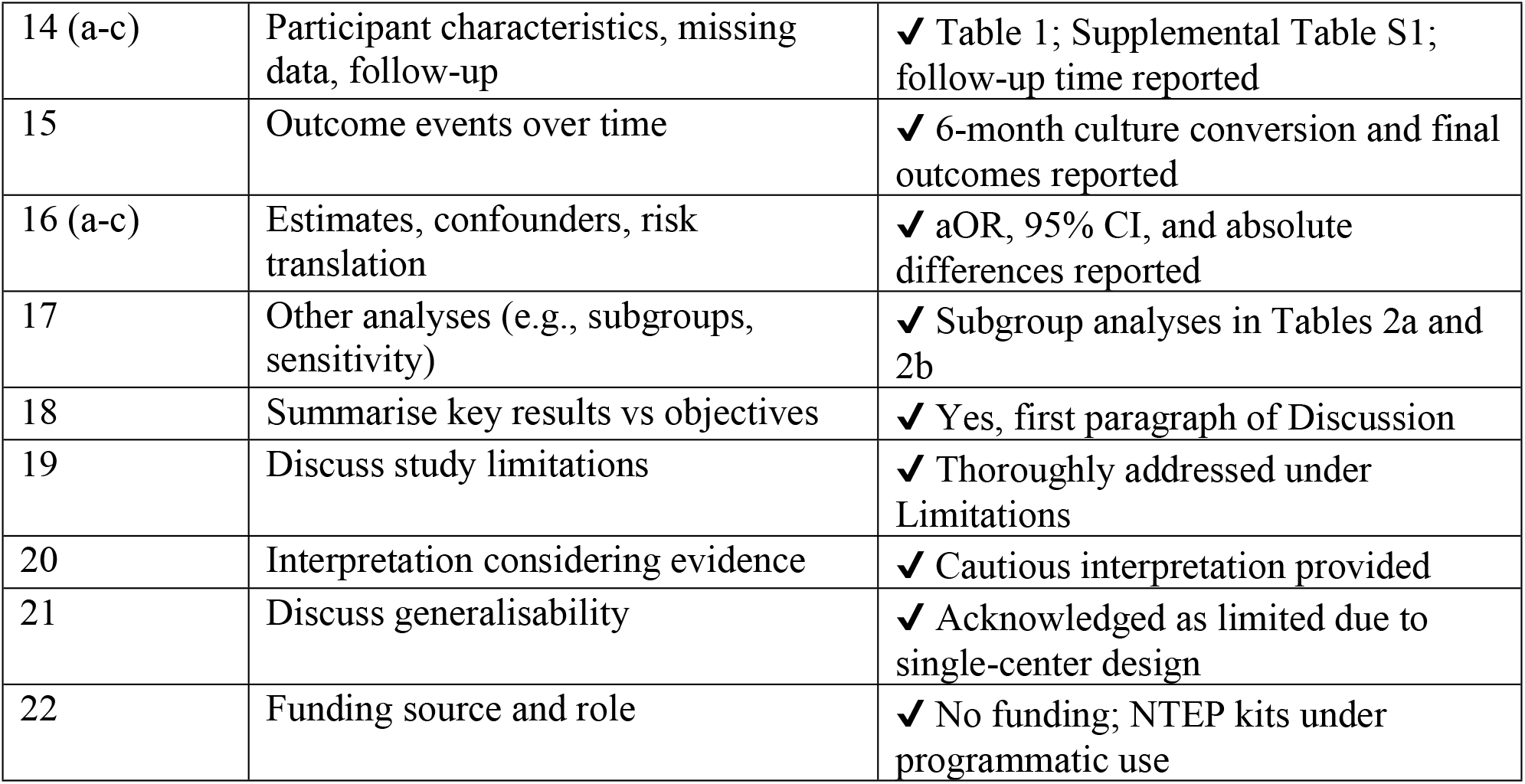

